# High performance of a novel antigen detection test on nasopharyngeal specimens for SARS-CoV-2 infection diagnosis: a prospective study

**DOI:** 10.1101/2020.10.28.20220657

**Authors:** L. Courtellemont, J. Guinard, C. Guillaume, S. Giaché, V. Rzepecki, A. Seve, C. Gubavu, K. Baud, C. Le Helloco, G.N. Cassuto, G. Pialoux, L. Hocqueloux, T. Prazuck

## Abstract

**Introduction:** The SARS-CoV-2 pandemic has become a major public health issue worldwide. Developing and evaluating rapid and easy-to-perform diagnostic tests is an absolute priority. The current study was designed to assess diagnostic performances of an antigen-based rapid detection test (COVID-VIRO®) in a real-life setting.

**Methods:** Two nasopharyngeal specimens of symptomatic or asymptomatic adult patients hospitalized in the Infectious Diseases Department or voluntarily accessing the COVID-19 Screening Department of the Regional Hospital of Orléans, France, were concurrently collected. COVID VIRO® diagnostic specificity and sensitivity were assessed in comparison to real-time reverse transcriptase quantitative polymerase chain reaction (RT-qPCR) results. A subset of patients underwent an additional oropharyngeal and/or a saliva swab for rapid testing.

**Results:** 121 patients already having a confirmed infection and 127 patients having no evidence of recent or ongoing infection were enrolled, for a total of 248 couple of nasopharyngeal swab specimens. Overall COVID-VIRO® sensitivity was 96.7% (IC: 93.5%-99.9%). In asymptomatic patients, symptomatic patients having symptoms for more than 4 days and those having a RT-qPCR Cycle threshold value ≥32, sensitivity was of 100%, 95.8% and 91.9% respectively. The concordance between RT-qPCR and COVID VIRO® rapid test was 100% for the 127 patients with no SARS-CoV-2 infection.

**Conclusion:** COVID-VIRO® test had 100% specificity and above 95% sensitivity, better than WHO recommendations (specificity ≥97-100%, sensitivity ≥80%). These rapid tests are particularly interesting for large-scale screening in Emergency Department, low resource settings and airports.

## Introduction

At the end of 2019, a pneumonia of initially unknown origin was primarily reported to the World Health Organization (WHO) Country Office in China. On January 9th, 2020, the Chinese health authorities and the WHO announced the discovery of a novel coronavirus, firstly named 2019-nCoV, then officially SARS-CoV-2 (Severe Acute Respiratory Syndrome CoronaVirus 2). This virus, belonging to the coronavirus family but differing from SARS-CoV-1 and MERSCoV, is responsible for upper/lower respiratory tract infections known as COVID-19 (COronaVIrus Disease 2019). COVID-19 incubation period is approximately 5.2 days and the most common onset symptoms are fever, cough, and fatigue (1). Since SARS-CoV-2 appeared in China, it has become a major public health issue worldwide. To date, more than 40 million cases have been detected worldwide (2) and the pandemic continues to spread unabated.

Minimizing testing delay seems to have the largest impact on reducing onward transmissions (3), and the availability of highly sensitive and specific tests is essential to quickly identify new cases and contain virus transmission.

Currently, the real-time reverse transcriptase quantitative polymerase chain reaction (RT-qPCR) assay is the gold-standard method to detect SARS-CoV-2 RNA in respiratory specimens such as nasopharyngeal or oropharyngeal swabs or broncho-alveolar lavage (4). However, performing RT-qPCR is expensive, time consuming, and requires special equipment and qualified operators. Faster, cheaper, and easier to use alternative tools could be represented by novels antigen-based rapid detection tests or point of care tests (POCTs) (5). Recently, the WHO approved the first rapid detection test for a large-scale use in low- and middle-income countries (6), and French health regulation authorities authorized their use in medical settings (7). Several different POCTs have already been developed (8), with generally high specificity but variable sensitivity (9-21).

COVID-VIRO® (AAZ, Boulogne Billancourt, France) is one of the novel immunochromatographic tests designed to detect SARS-CoV-2 antigen in nasopharyngeal specimens within 15 minutes (22). Evaluating diagnostic performances of COVID-VIRO® in the real life in comparison to the RT-qPCR as reference test is the principal aim of the current study.

## Materials and methods

### Ethical approval

This study was approved by the Regional North West Ethics and Research Committee. A written informed consent was obtained from each participant.

### Study population

People voluntarily accessing the COVID-19 Screening Department as well as subjects tested SARS-CoV-2 positive in the previous five days and SARS-CoV-2 positive patients hospitalized in the Infectious Diseases Department of the Centre Hospitalier Régional (CHR) of Orléans, France, or Drouot Laboratory, Paris, France, from October 12^th^, 2020, to October 25^th^, 2020, were included in the study. The diagnosis of SARS-CoV-2 infection was confirmed in case of positivity of the specific RT-qPCR on nasopharyngeal swab, in accordance with current recommendations. Subjects recently tested SARS-CoV-2 positive at the COVID-19 Screening Department were re-contacted and re-tested within five days. Inclusion criteria were age ≥ 18 years old and agreement to undergo two concurrent nasopharyngeal swabs for RT-qPCR and COVID-VIRO® analysis, respectively. Patient age was collected at inclusion, as well as symptom onset date for symptomatic patients. Suggestive symptoms were headache, fatigue, fever, or upper or lower respiratory symptoms. Asymptomatic patients were defined as those not reporting any of these symptoms.

### Case definition

SARS-CoV-2 positive subjects were either patients having a positive RT-qPCR at the time of inclusion if done in parallel with the rapid test, either patients having a previous positive RT-qPCR within five days and a positive or negative RT-qPCR at the time of study sampling.

SARS-CoV-2 negative subjects were patients having a negative RT-qPCR at the time of inclusion without any previous positive test.

### Specimen collection

Paired nasopharyngeal swabs were obtained for each patient by trained healthcare personnel (nurses, doctors, or biologists). The collection of the two simultaneous samples was always carried out by the same operator.

A polyester-tipped flexible (Viral transport medium tube with swab VTM, Sun-Trine^®^) was inserted into two of the nostrils until resistance was felt at the nasopharynx, then rotated 6 times and withdrawn. After swabbing, the swab applicator was cut off. The first absorbent swab was placed into a vial containing 3 mL of inactivating viral transport media and was immediately transferred to the Virology Unit of the CHR of Orléans hospital, Orléans, or Drouot Laboratory, Paris, to perform RT-qPCR. The rapid antigen test was immediately performed on-site with the second absorbent swab.

An additional oropharyngeal and/or saliva swab specimen was simultaneously collected in a subset of positive patients in order to determine the diagnostic reliability of these samples in comparison to nasopharyngeal swab specimens. Oropharyngeal specimens were collected on both sides of the tonsillar arches and posterior pharynx. Saliva specimens were collected by swabbing the upper and lower gingiva twice from back to front.

### Real-time RT-qPCR assays for the detection of SARS-CoV-2 RNA

Nucleic acid extraction was performed with automated Sample Preparation System MGISP-960 (MGI, China). Specific real-time RT-qPCR assays target three SARS-CoV-2 genes, namely ORF1ab, S and N genes (TaqPath Covid-19 Multiplex RT-PCR, Thermofisher). Genome amplification was performed using QuantStudio5 (Applied Biosystems). Results interpretation was performed according to manufacturer instructions. The assay includes an RNA internal extraction control and amplification control. Samples showing an exponential growth curve and a Cycle threshold (Ct) value < 37 were considered as positive. A Ct unique value > 37 was considered as negative.

### Rapid antigen test

COVID-VIRO® (AAZ, Boulogne Billancourt, France) is a membrane-based immunochromatography assay detecting SARS-CoV-2 nucleocapsid antigen (N-protein) in nasopharyngeal samples through monoclonal antibodies. A second monoclonal antibody is conjugate to colloidal gold particles which are captured in reaction membrane. The test was performed according to manufacturer instruction by mixing nasopharyngeal secretions with 300 µL of dilution buffer in a tube. One minute, 4 drops were added in the appropriate well. When nasopharyngeal secretions cross the strip, a passive diffusion allows the solubilized conjugate to migrate with the sample and react with the anti-SARS-CoV-2 antibodies immobilized on the membrane. A control line allows assessing the correct migration of sample and the reliability of the test. Visual interpretation of results is performed 15 minutes later (Figure 1).

**Figure 1:**
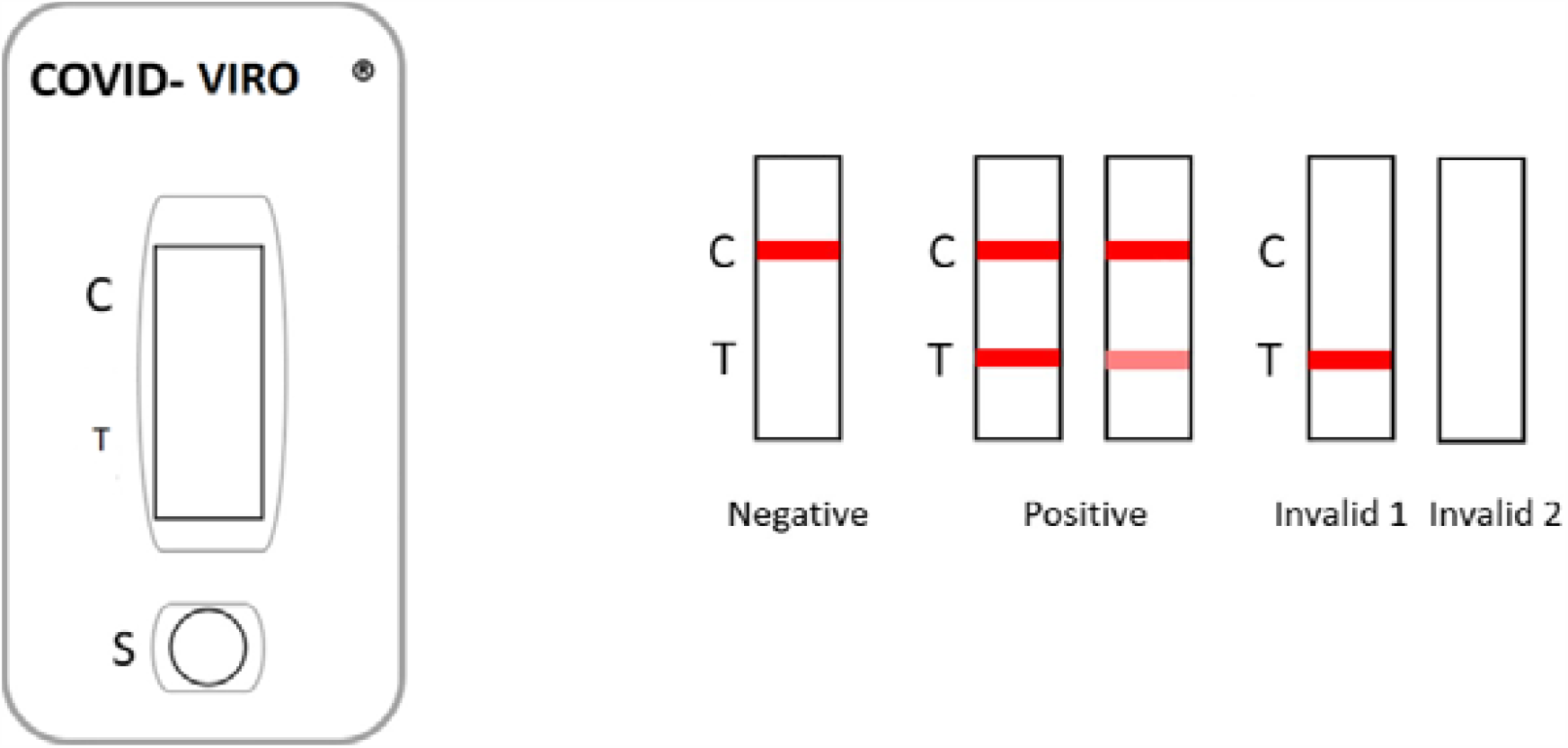
Interpretation of results for COVID-VIRO® (AAZ, LMB)

### Data analysis

Population characteristics are reported as percentage, mean and median values, standard deviation, and range. Data was analyzed in the Infectious Diseases Department.

To determine the diagnostic value of COVID-VIRO®, the study population was stratified into two groups:

1. Already confirmed RT-qPCR positive patients at the time of inclusion. Comparison between RT-qPCR and COVID-VIRO® results in these patients was used to assess diagnostic test sensitivity.
2. Non-selected symptomatic or asymptomatic patients voluntarily accessing the COVID-19 Screening Department in order to detect a possible SARS-CoV-2 infection. Analysing data, RT-qPCR positive patients were added to the first group in order to assess diagnostic test sensitivity. Conversely, RT-qPCR negative patients were selected to quantify the specificity of the rapid test.

COVID-VIRO® specificity and sensitivity were calculated using the RT-qPCR results as reference test, according to the following formulas:

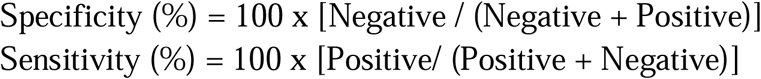

The size of the study population was calculated on the basis of a 95% sensitivity with a lower margin of the confidence interval over 91% and a 99% specificity with a lower margin over 95% according to the WHO recommendations for antigenic rapid test. T Student test was used to compare means.

## Results

121 patients with a SARS-CoV-2 confirmed infection and 127 patients with no evidence of recent or ongoing SARS-CoV-2 infection were enrolled in the study. A total of 248 couples of nasopharyngeal swabs were analyzed. Of these, 228 were collected in Orléans, and 20 in Paris.

The sex ratio of the study population was 0,9 (117 men and 131 women). The median and mean age was 38 and 43 years old, respectively (range: 18-96). Figure 2 shows the flow chart of included subjects in the study. One patient exhibiting only one positive target (gene S, Ct 36) at RT-qPCR analysis, was considered as negative according to the French Society of Microbiology criteria and the extraction kit instructions, and he was therefore included in the group of SARS-CoV-2 negative patients. 97 patients were symptomatic and 24 totally asymptomatic. No data about household or non-household close contacts were available.

**Figure 2:**
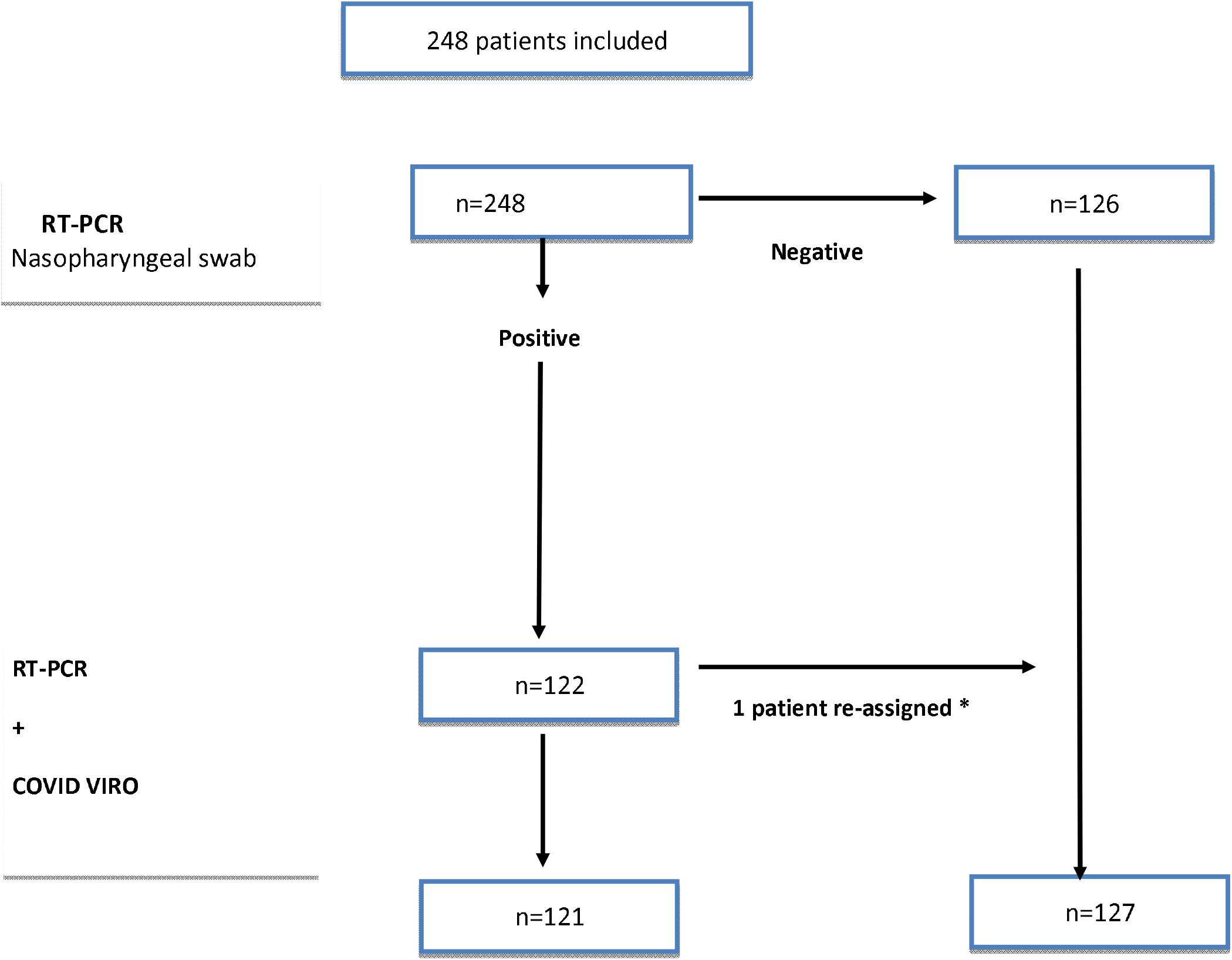
Flow Chart (*: one patient re-assigned, RT-PCR Ct >36 one target alone, false positive RT-qPCR)

Among the 121 patients diagnosed SARS-CoV-2 positive, 17 were hospitalized (14%) and 97 (80.1%) were symptomatic. The median time of symptoms duration before sampling was 5 days (mean: 6 days, range: 1-20).

N gene mean Ct value was 25 (range: 15-34) in asymptomatic SARS-CoV-2 positive patients, compared to 27 (range: 13-35) in symptomatic positive patients. The difference was not significant (p<0.05). In patients tested positive within 0 to 4 days from symptoms onset, the N gene mean Ct value was 25 (range: 13-35), versus 28 (range: 18-33) in those tested within 5 and 7 days, and 30 (range: 21-35) in those tested after more than 7 days, showing a constant decrease in viral carriage. Although some RT-qPCR false negative patients had a higher Ct value (suggestive of a lower RNA carriage), COVID-VIRO® was able to detect the antigen in 20 patients who had been reporting symptoms for more than 7 days.

Among the 121 RT-qPCR positive patients, 4 had a negative COVID-VIRO® result (3.3% false negative). The overall sensitivity of our POCT is estimated at 96.7% (IC: 93.5%-99.9%) (table 1). Among the 24 asymptomatic patients, no COVID-VIRO® false negative result was reported.

**Table 1:** Performances of the COVID-VIRO® antigenic rapid test in the overall population and in the group of asymptomatic SARS-CoV-2 infected patients

|  |  | COVID-VIRO® |  | COVID-VIRO® performances |  |
| --- | --- | --- | --- | --- | --- |
|  |  | Positive | Negative | Sensitivity(%<br>95% IC) | Specificity (%) |
| All patients | Positive RT-PCR | 117 | 4 | 96.7% (93.3-99.9%) |  |
|  | Negative RT-PCR | 0 | 127 |  | 100% |
| Asymptomatic<br>COVID-19 + patients | Positive RT-PCR | 24 | 0 | 100% |  |

Table 2 shows the COVID-VIRO® performances according to the Ct value and the delay of symptoms onset. COVID-VIRO® sensitivity was extremely high among patients having a Gene N, S or ORF Ct-values ≥32, considering that 34 out of 37 patients were tested positive sensitivity: 91.9% (95% IC: 83.1% - 100%).

**Table 2:**
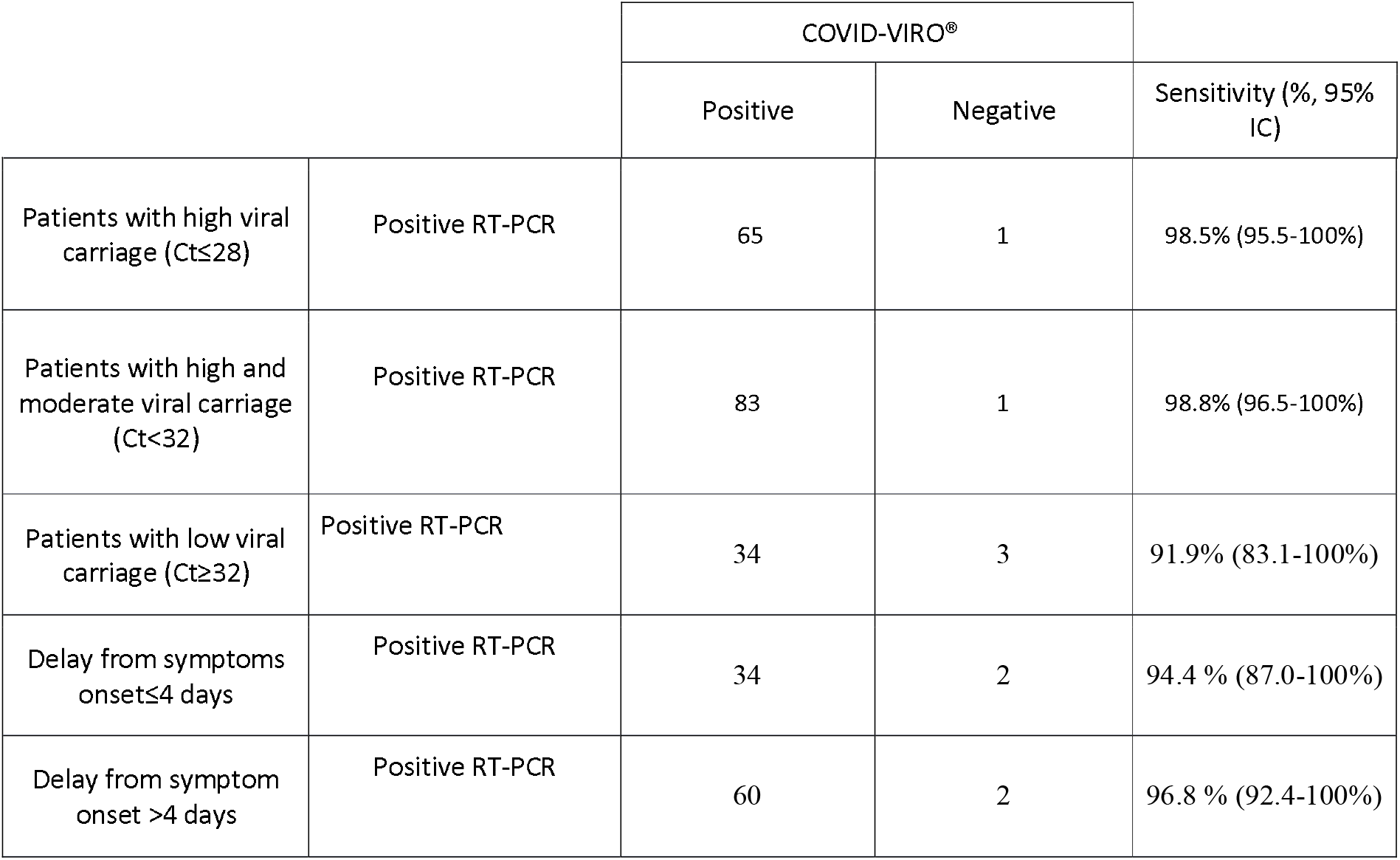
Sensitivity of the COVID-VIRO® antigenic rapid test in comparison to RT-PCR according to viral carriage and delay from symptom onset

Table 3 reports the characteristics of the four COVID-VIRO® false negative cases. Three out of four had Ct values ≥ 32 and were considered as non-contagious, regardless the date of onset of symptoms.

**Table 3:** Characteristics of the four discordant positives RT-PCR negative COVID-VIRO®. None of patient was hospitalized.

| Patient | Age range | Sex | Symptoms | S, N, ORF Gene Ct respectively | Delay from symptom onset |
| --- | --- | --- | --- | --- | --- |
| #1 | 20-30 | F | Yes | 31, 34, 31 | 5 |
| #2 | 20-30 | M | Yes | 32, 32, 31 | 1 |
| #3 | 60-70 | F | Yes | Neg, 32, 32 | 16 |
| #4 | 40-50 | M | Yes | 32, 28, 28 | 3 |

Twenty positive patients having a previous positive RT-qPCR were tested negative when the second RT-qPCR was simultaneously performed with the POCT. All of these had a positive COVID-VIRO® result (mainly weak or very weak line) suggesting that it may still be positive some days after the PCR negativization.

Among the 127 patients with no SARS-CoV-2 infection, no false positive result was observed as the concordance between RT-qPCR and COVID-VIRO® was 100%. COVID-VIRO® specificity is therefore estimated at 100%.

Additionally, 48 patients having a positive COVID-VIRO® test on nasopharyngeal swab specimen accepted undergoing a simultaneous oropharyngeal (34 patients) or saliva swab (14 patients). COVID-VIRO® turned positive in 24 out of 34 and 0 out of 14 patients on oropharyngeal and saliva specimens, respectively. Sensitivity was 70.6% for oropharyngeal and 0% for saliva specimens.

## Discussion

This observational study aims to evaluate the diagnostic performance of a POCT developed to detect SARS-CoV-2 antigens from a nasopharyngeal swab directly after sampling and provide the result within 15 minutes. The diagnostic value of COVID-VIRO® was determined using RT-qPCR as gold standard in a community setting. In the current study, COVID-VIRO® sensitivity and specificity were 96.7% and 100% respectively, with no observed false positive. Although results should be evaluated after 15 minutes, positive results almost always appeared within the first five minutes, not infrequently within one minute.

To date, several studies have evaluated the diagnostic performance of POCTs in real life, yielding conflicting data (9-21). In general, sensitivity ranged from 60.8% to 79.6% (13-18). Occasionally, significantly high sensitivity (93.9% and 98.3%) has been reported (19-21).

Concerning the Panbio COVID-19 Ag Rapid test (Abbott), the manufacturer reported a high sensitivity (93.3%; 95 CI 83.8-98.2) in a high endemic setting in Brazil (23) but other independent cohort studies have not confirmed these data. In 257 symptomatic and asymptomatic patients enrolled at the Emergency Department and Primary Health Care Setting in Spain, overall sensitivity was 73.3%, reaching 86.5% among patients having symptoms for less than seven days (11). In another multicentric study performed on 200 SARS-CoV-2 positive patients, POCT sensitivity was 72.6% (95% CI: 64.5-79.9%) in the Netherlands and 81.0% (95% CI: 69.0–89.8%) in Aruba. Test sensitivity was as high as 95.2% (95% CI: 89.3-98.5%) in patient with RT-qPCR test positivity to Ct-values < 32. (24).

In our study, stratifying patients by the Ct value, COVID-VIRO® sensitivity remained extremely high even for Ct values >32 (96.9%; 95% IC: 91.1% - 100%).

Detection of viral RNA on nasopharyngeal samples is not necessarily related to infectiousness (25). Several factors determine the risk of viral transmission: these include whether a virus is still viable, the amount of viral replication estimated by the Ct, the presence of respiratory symptoms, the individual’s local mucosal immune response to the virus, and the behaviour of the infected individual and their close contacts (26). However, in the present study the number of viral particles estimated by the Ct value did not differ in asymptomatic and symptomatic SARS-CoV-2 infected patients. COVID-VIRO® appears to be as sensitive as RT-qPCR to detect infected patients in a limited number of asymptomatic patients.

Available data report that RNA viral load rapidly decreases after the onset of symptoms, and infectiousness generally declines within 7–10 days (25, 27-30). Stratifying our patients by symptoms duration at sampling, we observed a similar decline from day 1 to day 14 in the mean Ct value enregistered. Only one hospitalized patient aged over 80, exhibited a Ct value of S :21, N: 22, ORF: 21 at day 10. Anyway, our POCT was able to detect the antigen both within and after 4 days since the onset of symptoms.

Considering discordant results, the analytical performances depend on different factors including viral load, quality of the specimen and processing. The two nasopharyngeal swabs were concurrently performed by the same operator, but a greater quantity of secretions and virus were likely to be more concentrated on the first one. Unfortunately, we cannot know the temporal order in which each swab was performed. Furthermore, waiting one hour between the two samples was infeasible.

Analysing POCT results on oropharyngeal and saliva specimens of positive patients, we quickly realized that sensitivity dramatically drops. Even if nasopharyngeal swab is uncomfortable for patients, this specimen should be privileged for the diagnosis of SARS-CoV-2 infection, whether obtained through PCR or POCTs.

This study has several limitations. First, this study is not a real-life study as when PCR positive patients came back for the study, the operators knew the status of the patient. Then, operators paid more attention to the results within 15 minutes. This method allows to give the real performances of the test done in the best conditions. This could be not the same in the real life, with many operators, emergencies ward where nurses will not necessary wait enough before reading or did not have the best conditions to read a thin line. Second, the date of symptoms onset was reported by patients and may not always be accurate, leading to an inaccurate stratification of patients. Then, the number of asymptomatic patients is rather limited to obtain conclusive data, even if we did not observe any discordance between tests in this group of patients. Finally, we did not perform any parallel comparison with other POCTs.

In France, nurses, pharmaceuticals, and general practitioners have recently been authorized to perform POCTs in medical settings (7). Data obtained from the current study could reassure health authorities in expanding access to POCTs that, in addition to being quick and easy to use, are also reliable. In the current setting, positive unaware people could wait for RT-qPCR results for several days. During this time, they could easily infect their close contacts. A more rapid diagnosis and the subsequent contact tracing would certainly positively impact on the containment of transmission.

In the Emergency Departments, POCTs could also be used to quickly recognise asymptomatic positive from negative patients, avoiding SARS-CoV-2 nosocomial infection. Furthermore, such tests would likely be useful in low- and middle-income countries and at airports to limit the viral spread worldwide.

## Conclusion

COVID-VIRO® (AAZ, Boulogne Billancourt, France) was a quite a reliable test for SARS-CoV-2 diagnosis. Diagnostic sensitivity and specificity on nasopharyngeal swabs were 96.7 % (95% CI 9 – 99.9%) and 100% respectively. To date, this is the unique COVID-19 antigenic rapid test fulfilling the WHO’s recommendations for a screening test (sensitivity ≥80%, specificity ≥97-100%). Unfortunately, performing the test on oropharyngeal or salivary samples is not as reliable.

## Data Availability

data are fully avalaible by request to the corresponding author

## Acknowledgement

The results of this study have already been mentioned by the manufacturer in the kit insert (22).

